# Equity analysis for Group-ANC in Ethiopia: Exploring how wealth and educational status interact with ANC attendance and facility delivery

**DOI:** 10.1101/2025.10.19.25338328

**Authors:** Ethan D Assefa, Solomon Getachew Alem, Walelegn Worku Yallew, Dedefo Teshite, Della Berhanu, Gadise Bekele, Stephanie Suhowatsky, Lisa Noguchi, Alemayehu Worku

**Author notes:** **Corresponding author:** (EDA). **Trial Registration**: ClinicalTrials.gov Identifier: NCT05054491.

## Abstract

**Background:** Group antenatal care promotes positive maternal health outcomes. We consider a health-equity perspective and ask whether group antenatal care at the health-post level in the rural Amhara region of Ethiopia can offer equal practicality and efficacy in elevating maternal health outcomes across socio-economic and educational levels.

**Methods:** Secondary analysis of a stepped-wedge trial was conducted in 36 health-posts in 2021-2023. Randomization of the order of the intervention introduction was done at the health-center level. Multivariable generalized estimating equation regression models were applied to analyze the two primary outcomes of at least four antenatal care visits and facility delivery, while exploring whether mother’s wealth level and mother’s education status as equity variables are associated with the outcomes through an interaction effect.

**Results:** Mothers in G-ANC in the middle wealth class had 82% lower odds of attending at least four ANC visits (OR= 0.18; 95% CI = [0.09, 0.34]; p <0.001), compared to the low wealth class. Mothers in G-ANC in the high wealth class had 92% lower odds of attending at least four ANC visits (OR= 0.08; 95% CI = [0.02, 0.32]; p <0.001), compared to the low wealth class. The intervention appeared more effective among mothers with formal education, who had over twice the odds of completing ANC visits compared to their counterparts with no schooling, though the association was marginally non-significant (OR = 2.36; 95% CI= [0.98–5.70]; p = 0.057). The interactions between the intervention and equity variables for facility delivery were non-significant.

**Conclusion:** Poorer mothers in G-ANC significantly benefitted more than those in higher wealth classes. It was less effective among mothers with no education, but not statistically significant. No effect was found of wealth or education on facility delivery.

## 1. Introduction

Wealth and education influence antenatal care (ANC) use in low- and middle-income countries (LMICs). Lower-income and lower-educated women have reduced odds of attending at least four antenatal care (ANC4+) visits [1–4], including in Ethiopia [5]. Rural residence has also been similarly associated with lower ANC utilization [6,7]. This evidence points to the importance of ensuring equitable access to healthcare during pregnancy and childbirth considering utilization of maternal health care services is key to improving health outcomes [8–10].

Group antenatal care (G-ANC) is an alternative care model to the current standard of care, individual ANC (I-ANC). Group antenatal care (G-ANC) is an intervention where pregnant women at similar stages of pregnancy are assigned to cohorts and receive antenatal care together in group sessions. These sessions typically include clinical assessments alongside structured group discussions and activities, facilitated by healthcare providers, such as Health Extension Workers (HEWs) in the context of Ethiopia. This is opposed to Individual antenatal care (I-ANC), which refers to the conventional standard of care where pregnant women receive antenatal care on a one-to-one basis with a healthcare provider. In high-income countries, G-ANC has been shown to improve adherence, satisfaction, and health outcomes for the expectant mothers and newborns [11]. The World Health Organization (WHO) in 2016 recommended G-ANC as a health system intervention in the context of rigorous research [12]. Since then, a number of studies in LMICs have shown G-ANC to be an effective intervention [13–15], at both individual and systemic levels. G-ANC is positively associated with increased ANC coverage [16–19], postnatal care use [16–18,20], improved quality of care [16,18,21,22] and use of postpartum family planning (PPFP) [23,24] when compared with I-ANC. G-ANC is also associated with increased facility delivery in Nigeria and Ethiopia [16,25]. Additionally, women in G-ANC reported greater satisfaction [16,17,21,26,27], as well as increased feelings of pregnancy-related empowerment and self-efficacy [18,28] when compared with those in I-ANC.

A research trial was conducted in the rural Amhara region to introduce G-ANC at the health post level. The health post serves as the lowest level of primary care in Ethiopia. It is staffed with two professionals Health Extension Workers (HEWs) who know their local community well and are focused on maternal and child health [29–31]. Our trial reported G-ANC was acceptable, effective, and feasible at the health posts, and the G-ANC intervention was found to significantly increase odds of ANC4+ visits and facility delivery [25]. This study examines how participants’ wealth and education interact with G-ANC at the health post level to influence ANC attendance and delivery at a health facility.

While it has been shown that G-ANC is an effective and feasible solution to improve maternal health outcomes for provincial Amhara regions at the health post level across the general population, it is not known that it remains effective and feasible for vulnerable populations, such as those in lower wealth classes and the less-educated, in these same rural Amhara regions of Ethiopia. Our work addresses this gap and, in doing so, examines this issue in two ways. First, with a clinical trials lens, we utilize the data from a stepped-wedge design with randomization of intervention order and comparisons across arms. Second, with a health equity lens, we explore whether vulnerable groups, particularly the poorest and less-educated, receive the same beneficial impact of G-ANC, compared with the more privileged participants.

## 2. Materials and Methods

### 2.1 Data Collection and Study Design

This study was secondary analysis of data collected by Addis Continental Institute of Public Health (ACIPH) and Jhpiego for assessing effectiveness, acceptability and feasibility of G-ANC at the health post level in the rural Amhara region in Ethiopia [25]. Data from this study, collected in 2022-2023, were utilized as the starting point for our analysis. The trial consisted of a stepped-wedge design [32] conducted in 36 health posts under the umbrella of six health centers. The order in which the intervention was introduced occurred at the health-center level in a cluster randomization design. The study intended to have three time periods: an initial period of six months with no G-ANC, followed by six months where G-ANC was introduced in half (n = 18) of the health posts. Then, in the final six months, G-ANC was supposed to be implemented in the remaining 18 health posts. However, only data from the first two periods were able to be collected, due to local security conditions and safety concerns. Details on data collection methodology are available in the primary study’s methods section [25].

The original trial was approved by the ethics committee of the Addis Continental Institute of Public Health Institutional Ethical Review Board (ACIPH/IRB/006/2021) in Addis Ababa, Ethiopia, on June 16, 2021, and the Johns Hopkins Bloomberg School of Public Health Institutional Review Board (IRB14448/284) in Baltimore, Maryland, USA, on September 10, 2021. Research assistants obtained informed consent from women in a private space using IRB-approved recruitment and consent documents in the local language. Verbal consent was preferred over written consent to respect community norms and ensure confidentiality. The present secondary analysis collected no new data, involved no additional contact with participants, and adhered to the ethical standards set forth in the initial trial. Data for this secondary analysis were accessed on November 19, 2024, following a formal data-sharing agreement. The dataset contained no personally-identifying information. Variables with potential indirect identifiers, such as health post or health center, were recoded into numerical codes for clustering purposes only. All data were handled securely and in accordance with the confidentiality protections approved in the original trial protocol.

### 2.2 Analysis

Exploratory data analysis examined our overall sample, and descriptive statistics were calculated for the distributions of the variables of interest (mean and SD for continuous variables, counts and percentages for categorical). Missing data were minimal, with ∼99.5% of the total data present (see Appendix Table S1 for missingness). Key variables of interest—their name, purpose, data type, and description—are listed below.

**Table 1.** Study variables of interest for equity analysis from the G-ANC trial in health posts in Amhara Region, Ethiopia.

| Name | Purpose | Type | Description |
| --- | --- | --- | --- |
| Participant Study ID | Identity | Numeric | Participant identifier code assigned to individuals when entering the study. |
| Health Post Code | Grouping | Categorical | Health post identifier code assigned to each respective health post. Names of health facilities were left unextracted as they were unnecessary for analysis and to preserve ethics. |
| Health Center Code | Grouping | Categorical | Health center identifier code assigned to each respective health center, under which the health posts were grouped. Names of health facilities were left unextracted as they were unnecessary for analysis and to preserve ethics. |
| Intervention Status | Intervention | Binary | Participant's randomization group, informing whether they were assigned to the intervention or control group. |
| Age Group | Demographic | Categorical | Participant's age; defined as categories 15-24 years old, 25-34 years old, and 35-49 years old. |
| Education Status | Equity | Binary | Participant's educational status, defined as having ever/never attended school. |
| Wealth Quintile | Equity | Categorical | Wealth index score distribution of participants grouped into 20% quintiles. |
| Wealth Index Raw Score [33,34] | Equity | Numeric | Wealth index score as a continuous measure, calculated using the revised 2018 approach based on the Ethiopia DHS 2016 Equity Tool. |
| Wealth Category [33,34] | Equity | Categorical | Wealth categories created by collapsing quintiles into 3-level categories, based on DHS-defined methodology [33,34]; low is defined as the poorest 40%, middle is defined as the center 20%, high is defined as the richest 40%. |
| Distance from Health Post | Demographic | Numeric | Distance of the participant from their nearest health post, in minutes of travel. |
| Binary Distance Groups [35] | Equity | Binary | Distance binary created by splitting distance, based off previous work [35]; near was defined as less than or equal to 45 minutes, and far was defined as greater than 45 minutes. |
| ANC4+ Visits | Outcome | Binary | Binary variable indicating whether participant has attended at least four antenatal care visits (ANC) during the study period or not. |
| Place of Delivery | Outcome | Binary | Binary variable indicating whether participant has given birth in a health facility or not. |

We employed a difference-in-differences approach to assess whether the effect of G-ANC on selected equity variables varied across groups. We applied Generalized Estimating Equations (GEE) to account for site-level bias stemming from the clustering of data across different health centers [36,37]. We focus on education level (mother has attended school/mother has never attended school) and wealth level (low: bottom 40%, middle: center 20%, high: top 40%) as our equity group variables. The wealth score was calculated from utilizing the revised approach based on the Ethiopia Demographics and Health Survey (DHS) 2016 Equity Tool update released Jan 5, 2018, involving a simplified list of 15 equity-related questions measuring relative wealth specific to the nation and urbanicity of the individual [33]. The interaction between our equity group variables and the study intervention variable (whether the mother was assigned to G-ANC) were assessed through our models. The two primary outcomes from the trial are also the outcomes of interest here: whether the mother attended at least four ANC visits (ANC4+), and whether the mother had a facility delivery. The health center covering the health post where the mother received ANC was used as the clustering variable for our GEE models. Odds ratios were calculated and are presented in the Results section below. We applied a significance level of 0.05 for our p-values and reported 95% Confidence Intervals (CI) along with the point estimates.

This analysis was conducted with R version 4.4.2. The name, version, and description of all the R packages used are included in Table S2 in the appendix.

## 3. Results

### 3.1 Sample Demographics

Table 2 below shows general distributions of participants and the variables of interest across our study sample. There was a total of 537 participants, with 358 randomized to the control group and 179 randomized to the intervention. No significant difference in demographic/equity variables was noted between the intervention and control groups for age, education, or distance from the health post were found. Wealth was the only significant variable, both overall for the raw wealth index score (p <0.001) and the 3-level wealth categories (p = 0.002). There were a higher proportion of low-wealth status participants in the intervention group, compared to the other two wealth categories. Meanwhile, there is a larger proportion of high-wealth status participants in the control group, compared to the other two wealth categories. Tables S3 and S4 (see appendix) contain the demographics distributions stratified by wealth class groups and education status, respectively.

**Table 2.** Equity Analysis of the G-ANC trial in health posts in Amhara Region, Ethiopia - Overall study sample characteristics.

| Characteristic | Overall N = 537<sup>1</sup> | Control N = 358<sup>1</sup> | Intervention N = 179<sup>1</sup> | p-value<sup>2</sup> |
| --- | --- | --- | --- | --- |
| **Age Group** |  |  |  | 0.052 |
| 15-24 | 137 (26%) | 97 (27%) | 40 (22%) |  |
| 25-34 | 266 (50%) | 183 (51%) | 83 (46%) |  |
| 35-49 | 134 (25%) | 78 (22%) | 56 (31%) |  |
| **Education Status** |  |  |  | 0.11 |
| Mother never attended school | 277 (52%) | 176 (49%) | 101 (56%) |  |
| Mother has attended school | 260 (48%) | 182 (51%) | 78 (44%) |  |
| **Wealth Index Raw Score** | -0.33 (0.28) | -0.31 (0.27) | -0.39 (0.28) | <0.001 |
| **Wealth Category** |  |  |  | 0.002 |
| Low | 217 (40%) | 126 (35%) | 91 (51%) |  |
| Middle | 105 (20%) | 75 (21%) | 30 (17%) |  |
| High | 215 (40%) | 157 (44%) | 58 (32%) |  |
| **Distance from Health Post** | 34.34 (31.48) | 34.97 (32.24) | 33.07 (29.95) | 0.8 |
| **Binary Distance Group** |  |  |  | 0.2 |
| Far | 126 (23%) | 90 (25%) | 36 (20%) |  |
| Near | 411 (77%) | 268 (75%) | 143 (80%) |  |

<sup>1</sup>n (%); Mean (SD)
<sup>2</sup>Pearson's Chi-squared test; Wilcoxon rank sum test

### 3.2 Regression Analysis

Table 3 presents the results from the GEE regression model examining the interaction between intervention and wealth status. For the outcome of ANC4+ visits, we see significant interaction effects between assignment to the intervention and both the middle-income group (OR= 0.18; 95% CI = [0.09, 0.34]; p < 0.001) and high-income group (OR= 0.08; 95% CI = [0.02, 0.32]; p < 0.001). Those of middle- and high-income status in the intervention group were found to have lower odds of ANC4+ visits compared to those of low income. Significant associations were seen for the interaction of intervention and wealth status, which were not seen for the wealth categories on their own.

For the outcome of having a facility delivery, there was no statistically significant interaction effect between assignment to the intervention and the mother being middle income (OR= 0.64; 95% CI = [0.19, 2.21]; p = 0.5) or high income (OR= 1.10; 95% CI = [0.46, 2.61]; p = 0.8).

**Table 3.** GEE regression models on intervention-wealth interaction of the G-ANC trial in health posts in Amhara Region, Ethiopia.

| Characteristic | ANC4+ Visits |  |  | Facility Delivery |  |  |
| --- | --- | --- | --- | --- | --- | --- |
|  | OR<sup>1</sup> | 95% CI<sup>1</sup> | p-value | OR<sup>1</sup> | 95% CI<sup>1</sup> | p-value |
| Intervention Status |  |  |  |  |  |  |
| Control | — | — |  | — | — |  |
| Intervention | 90.4 | 11.3, 723 | <0.001 | 2.18 | 1.05, 4.53 | 0.037 |
| Wealth Category |  |  |  |  |  |  |
| Low | — | — |  | — | — |  |
| Middle | 0.84 | 0.57, 1.23 | 0.4 | 1.15 | 0.68, 1.94 | 0.6 |
| High | 1.07 | 0.65, 1.75 | 0.8 | 2.10 | 1.11, 4.00 | 0.023 |
| Age Group |  |  |  |  |  |  |
| 15-24 | — | — |  | — | — |  |
| 25-34 | 1.47 | 1.10, 1.96 | 0.010 | 0.73 | 0.43, 1.23 | 0.2 |
| 35-49 | 1.20 | 0.62, 2.34 | 0.6 | 0.72 | 0.41, 1.27 | 0.3 |
| Binary Distance Group |  |  |  |  |  |  |
| Far | — | — |  | — | — |  |
| Near | 1.17 | 0.79, 1.72 | 0.4 | 0.94 | 0.61, 1.44 | 0.8 |
| Education Status |  |  |  |  |  |  |
| Mother never attended school | — | — |  | — | — |  |
| Mother has attended school | 1.30 | 0.86, 1.97 | 0.2 | 1.39 | 0.91, 2.13 | 0.13 |
| Intervention Status * Wealth Category |  |  |  |  |  |  |
| Intervention * Middle | 0.18 | 0.09, 0.34 | <0.001 | 0.64 | 0.19, 2.21 | 0.5 |
| Intervention * High | 0.08 | 0.02, 0.32 | <0.001 | 1.10 | 0.46, 2.61 | 0.8 |

<sup>1</sup>OR = Odds Ratio, CI = Confidence Interval

Table 4 presents the GEE regression analysis on the interaction between intervention and educational status. For the outcome of attending ANC4+ visits, there is a marginally non-significant interaction effect between assignment to the intervention and the mother having attended school (OR= 2.36; 95% CI = [0.98, 5.70]; p = 0.057). The notably increased odds of ANC4+ visits are seen for the interaction of the two, even as the education status predictor on its own had a much smaller, non-significant difference (OR= 1.26; 95% CI = [0.80, 1.97]; p = 0.3). As in the primary study, the assignment to the intervention on its own was found to significantly increase odds of attending ANC4+ visits (OR= 18.7; 95% CI = [4.84, 72.0]; p < 0.001).

For the outcome of having a facility delivery, there was no significant interaction effect between assignment to the intervention and the mother having attended school (OR= 1.22; 95% CI = [0.61, 2.47]; p = 0.6).

**Table 4.** GEE regression models on intervention-education interaction of the G-ANC trial in health posts in Amhara Region, Ethiopia.

| Characteristic | ANC4+ Visits |  |  | Facility Delivery |  |  |
| --- | --- | --- | --- | --- | --- | --- |
|  | OR<sup>1</sup> | 95% CI<sup>1</sup> | p-value | OR<sup>1</sup> | 95% CI<sup>1</sup> | p-value |
| Intervention Status |  |  |  |  |  |  |
| Control | — | — |  | — | — |  |
| Intervention | 18.7 | 4.84, 72.0 | <0.001 | 1.83 | 0.79, 4.21 | 0.2 |
| Education Status |  |  |  |  |  |  |
| Mother never attended school | — | — |  | — | — |  |
| Mother has attended school | 1.26 | 0.80, 1.97 | 0.3 | 1.30 | 0.80, 2.12 | 0.3 |
| Age Group |  |  |  |  |  |  |
| 15-24 | — | — |  | — | — |  |
| 25-34 | 1.47 | 1.09, 1.98 | 0.011 | 0.72 | 0.42, 1.22 | 0.2 |
| 35-49 | 1.20 | 0.61, 2.37 | 0.6 | 0.72 | 0.42, 1.24 | 0.2 |
| Binary Distance Group |  |  |  |  |  |  |
| Far | — | — |  | — | — |  |
| Near | 1.15 | 0.78, 1.68 | 0.5 | 0.92 | 0.59, 1.45 | 0.7 |
| Wealth Category |  |  |  |  |  |  |
| Low | — | — |  | — | — |  |
| Middle | 0.76 | 0.51, 1.15 | 0.2 | 1.00 | 0.60, 1.68 | >0.9 |
| High | 0.95 | 0.57, 1.59 | 0.8 | 2.11 | 1.25, 3.55 | 0.005 |
| Intervention Status * Education Status |  |  |  |  |  |  |
| Intervention * Mother has attended school | 2.36 | 0.98, 5.70 | 0.057 | 1.22 | 0.61, 2.47 | 0.6 |

<sup>1</sup>OR = Odds Ratio, CI = Confidence Interval

## 4. Discussion

### 4.1 Key Interpretations

The association of the G-ANC intervention with completing ANC4+ visits has about 82% weaker odds for the middle-income group and about 92% weaker odds for the high-income group, both relative to the low-income group. This information, along with our findings of progressively reductive interaction effect as wealth increases compared to the low-income group, has interesting implications. It may point to the G-ANC intervention having little effect on those of higher wealth because the wealthiest mothers in both treatment groups, G-ANC and control, already have high utilization of ANC services and so there is no meaningful difference found.

This suggests G-ANC as a feasible intervention at the HP-level in Ethiopia, which can be used as an equity strategy to increase ANC attendance among low-income populations [38]. Women who were educated and in the intervention group were twice as likely to complete ANC4+ visits (OR= 2.36; 95% CI = [0.98, 5.70]; p = 0.057), compared to those with no education. While this finding was non-significant, it may suggest a unique modulating effect of a mother’s education status on the effectiveness of the intervention and further work may serve to elucidate this relationship.

Unlike its clear influence on ANC attendance, the G-ANC intervention did not show a significant differential effect on facility-based delivery across wealth or education subgroups. One explanation may be that women face different or persistent barriers to delivering in a health facility, which are not addressed by the G-ANC model itself. Studies in sub-Saharan Africa have documented that many women who obtain ANC still end up delivering at home due to factors such as distance to health centers, lack of transportation, cost concerns, and deeply rooted cultural norms [39]. Qualitative evidence from the Ethiopian context confirms a range of deterrents to facility delivery, including sudden labor onset at night, reliance on traditional birth attendants, privacy concerns, and family desires to be present during home birth [40]. It is plausible that G-ANC, while increasing engagement during pregnancy, did not mitigate these barriers to institutional delivery. Additionally, if both intervention and control groups were exposed to parallel initiatives promoting facility births (for example, Ethiopia’s free maternity services and community advocacy), the marginal added effect of G-ANC on delivery location may have been minimal. Dedicated measures such as birth preparedness counseling, transport support, or maternity waiting homes might be needed alongside G-ANC to ensure more women deliver in health facilities.

Global evidence consistently shows large wealth-based disparities in ANC utilization [1–5,41]. Against this backdrop, prior evaluations of G-ANC in LMICs have demonstrated overall improvements in ANC attendance and quality for the general population [16–19], but have not examined whether these benefits differ across socio-economic groups. Our finding that the G-ANC effect was strongest among low-income women is a novel contribution to the literature, suggesting that G-ANC may serve as an equity-promoting intervention. In other words, by markedly boosting ANC attendance among the most disadvantaged while having less influence on already high-utilizing affluent groups, G-ANC could help narrow the wealth gap in ANC service use. Efforts to leverage G-ANC as a means to improve equity have also been explored in high-income settings. A recent trial in the United States found that while overall birth outcomes were similar between care models, Black women in G-ANC had infants with rates of low birthweight closer to their White counterparts, effectively reducing the racial disparity [43]. To our knowledge, our study is among the first in a low-income country to demonstrate the equity impact of G-ANC, underscoring the potential of G-ANC to advance health equity in ANC utilization.

Historically, certain groups have been marginalized and overlooked in medical research and public health interventions. Even as the field of public health commits to better addressing such inequities [44,45], we continue to see their lasting effects in the health disparities present today [46–49]. In this study, an intervention shown to be effective generally was examined to see if it will hold up when examined for its effect across wealth and educational status. We saw how differences in efficacy can be seen for these vulnerable groups.

The notably stronger impact of G-ANC on ANC attendance among low-income women suggests this model can help bridge longstanding disparities in maternal care access. In Ethiopia, wealth-based gaps in ANC utilization are stark, with the poorest women 43 percent less likely to receive ANC4+ visits than the least-poor [50]. Addressing such inequities is a national priority and reducing socio-economic disparities in maternal health service coverage is explicitly highlighted as a key strategy in Ethiopia’s health sector plans [50]. Our finding that G-ANC most benefited women of lowest wealth directly align with these policy goals. By boosting ANC uptake in disadvantaged groups, G-ANC serves as a potential equity-promoting intervention that supports the Federal Ministry of Health’s focus on “quality and equitable ANC” services [51]. In practical terms, implementing G-ANC at the community level (health posts) could complement Ethiopia’s existing strategies, such as the Health Extension Program [52,53] and Health Equity Strategy [54,55], which strive to expand ANC access for underserved rural and low-income populations.

### 4.2 Recommendations

Our results point out several promising directions for future research. First, an expanded implementation of G-ANC at the health-post level is recommended to verify that the benefits observed in this trial are replicable at scale. Conducting a larger roll-out, covering more areas or regions outside the Amhara region, would allow for a robust evaluation of whether G-ANC consistently improves ANC uptake among the poorest women and truly narrows equity gaps in diverse settings.

Additionally, ongoing research should continue to incorporate an equity lens in intervention design – routinely stratifying outcomes by socio-economic and other vulnerable group markers – so that maternal health innovations such as G-ANC can be refined to maximize impact on those who need them most. Aggregate statistics and single axes approaches can, at times, overlook key groups. Interventions that do not consider and account for these identities will struggle to retain their efficacy and impact within these segments of the population [56–58]. Unfortunately, there remains a lack of consensus in scientific literature as to considering and operationalizing intersectionality when designing intersectional studies. More work needs to be done to ensure interventional studies routinely ponder the differential effects of their treatment across special groups, not only in analysis but also in design [59,60].

With around 15,000 health posts throughout the nation and a sizeable portion of the population with little education and living at the poverty level [61–64], our work elucidating the effectiveness of G-ANC at this level for these population segments has the potential to promote wide-ranging benefits for maternal and neonatal health across the country. Using our work, in conjunction with the general body of work on G-ANC, policymakers should tailor their planned interventions and large-scale government programs to best target not only the privileged, but also economically disadvantaged and uneducated women.

### 4.3 Limitations

This study contains limitations. As a secondary analysis, the data we analyzed was already collected at the time of the study proposal. The primary study’s data collection phase was prematurely ended due to security concerns and issues of data collector safety in the study area. However, the first two completed phases taken together include both control and intervention groups. Having found statistically significant interaction effects even without the full number of intervention and controls, we would expect increased sample group size to have only increased the study’s power and the resulting significance of our associations.

## 5. Conclusion

Significant wealth, but not education, differentials were found for ANC attendance among G-ANC participants. The poorest mothers benefitted more than those of higher wealth status, which is encouraging from a health-equity perspective as it narrows utilization gaps and inequity in care. There was a marginally non-significant effect for those who have lower education status.

Facility delivery didn’t see any significant differences by wealth or education. G-ANC at the health post level in Ethiopia appears a viable equity strategy for vulnerable and oft-overlooked populations.

## Data Availability

The fully deidentified data underlying the results presented in the study are available from Figshare at the following URL. URL: https://figshare.com/s/d49bb543396ba860929c

## 6. Acknowledgments

## 6.1 Authors’ contributions

WWY, DT, DB, SS, LN and AW contributed to the conceptualization of the primary study. EDA and AW analyzed, interpreted the data, and drafted the manuscript. SGA, WWY, DT, DB, GB, SS, and LN reviewed and commented on the manuscript. All authors have read and approved the final manuscript.

## A. Appendix

**Table S1.** Data/variable missingness counts in the equity analysis for the Group-ANC intervention in Ethiopia.

| Variable Name | Description | Missing Count | Missing Percentage* |
| --- | --- | --- | --- |
| Place of Delivery | Binary variable indicating whether participant has given birth to their baby in a health facility or not. | 32 | 5.96% |
| ANC4+ Visits | Binary variable indicating whether participant has attended at least four or more antenatal care visits (ANC) during the study period or not. | 6 | 1.12% |
| **All other variables have no missing observations** |  |  |  |

\* Percentage calculated as proportion of observations for the variable, not entire dataset

**Table S2.** R packages used in equity analysis for the Group-ANC intervention in Ethiopia.

| Package | Version | Description |
| --- | --- | --- |
| **tidyverse** | 2.0.0 | Primary data science package for R |
| **haven** | 2.5.4 | Reading in dta files |
| **labelled** | 2.13.0 | Converting number codes in dta metadata to labels |
| **naniar** | 1.1.0 | Visualizing missingness |
| **flipTime** | 2.10.0 | Converting dates/times to appropriate data type |
| **gtsummary** | 2.0.4 | Creating publication-ready tables |
| **geepack** | 1.3.12 | Provides Generalized Estimating Equation (GEE) functionality |

**Table S3.** Equity Analysis of the G-ANC trial in health posts in Amhara Region, Ethiopia - Study sample characteristics across wealth status.

| Characteristic | Low |  |  |  | Middle |  |  |  | High |  |  |  |
| --- | --- | --- | --- | --- | --- | --- | --- | --- | --- | --- | --- | --- |
|  | Overall<br>N =<br>217<sup>1</sup> | Control<br>N =<br>126<sup>1</sup> | Intervention<br>N = 91<sup>1</sup> | p-value<sup>2</sup> | Overall<br>N =<br>105<sup>1</sup> | Control<br>N = 75<sup>1</sup> | Intervention<br>N = 30<sup>1</sup> | p-value<sup>2</sup> | Overall<br>N =<br>215<sup>1</sup> | Control<br>N =<br>157<sup>1</sup> | Intervention<br>N = 58<sup>1</sup> | p-value<sup>2</sup> |
| **Age Group** |  |  |  | 0.082 |  |  |  | 0.5 |  |  |  | 0.078 |
| 15-24 | 54<br>(25%) | 35<br>(28%) | 19 (21%) |  | 27<br>(26%) | 20<br>(27%) | 7 (23%) |  | 56<br>(26%) | 42<br>(27%) | 14 (24%) |  |
| 25-34 | 108<br>(50%) | 66<br>(52%) | 42 (46%) |  | 47<br>(45%) | 31<br>(41%) | 16 (53%) |  | 111<br>(52%) | 86<br>(55%) | 25 (43%) |  |
| 35-49 | 55<br>(25%) | 25<br>(20%) | 30 (33%) |  | 31<br>(30%) | 24<br>(32%) | 7 (23%) |  | 48<br>(22%) | 29<br>(18%) | 19 (33%) |  |
| **Education Status** |  |  |  | 0.4 |  |  |  | 0.6 |  |  |  | 0.2 |
| Mother never attended school | 126<br>(58%) | 70<br>(56%) | 56 (62%) |  | 60<br>(57%) | 44<br>(59%) | 16 (53%) |  | 91<br>(42%) | 62<br>(39%) | 29 (50%) |  |
| Mother has attended school | 91<br>(42%) | 56<br>(44%) | 35 (38%) |  | 45<br>(43%) | 31<br>(41%) | 14 (47%) |  | 124<br>(58%) | 95<br>(61%) | 29 (50%) |  |
| **Distance from Health Post** | 38.40<br>(32.62) | 37.78<br>(32.55) | 39.25<br>(32.87) | 0.6 | 34.62<br>(28.44) | 38.81<br>(31.13) | 24.13<br>(16.42) | 0.048 | 30.10<br>(31.31) | 30.87<br>(32.24) | 28.00<br>(28.78) | 0.7 |
| **Binary Distance Group** |  |  |  | 0.9 |  |  |  | 0.007 |  |  |  | 0.2 |
| Far | 63<br>(29%) | 36<br>(29%) | 27 (30%) |  | 26<br>(25%) | 24<br>(32%) | 2 (6.7%) |  | 37<br>(17%) | 30<br>(19%) | 7 (12%) |  |
| Near | 154<br>(71%) | 90<br>(71%) | 64 (70%) |  | 79<br>(75%) | 51<br>(68%) | 28 (93%) |  | 178<br>(83%) | 127<br>(81%) | 51 (88%) |  |

<sup>1</sup>n (%); Mean (SD)
<sup>2</sup>Pearson's Chi-squared test; Wilcoxon rank sum test

**Table S4.** Equity Analysis of the G-ANC trial in health posts in Amhara Region, Ethiopia - Study sample characteristics across education status.

| Characteristic | Mother never attended school |  |  |  | Mother has attended school |  |  |  |
| --- | --- | --- | --- | --- | --- | --- | --- | --- |
|  | Overall N<br>= 277<sup>1</sup> | Control N<br>= 176<sup>1</sup> | Intervention N<br>= 101<sup>1</sup> | p-value<sup>2</sup> | Overall N<br>= 260<sup>1</sup> | Control N<br>= 182<sup>1</sup> | Intervention N<br>= 78<sup>1</sup> | p-value<sup>2</sup> |
| **Age Group** |  |  |  | 0.5 |  |  |  | 0.093 |
| 15-24 | 26 (9.4%) | 16 (9.1%) | 10 (9.9%) |  | 111 (43%) | 81 (45%) | 30 (38%) |  |
| 25-34 | 134 (48%) | 90 (51%) | 44 (44%) |  | 132 (51%) | 93 (51%) | 39 (50%) |  |
| 35-49 | 117 (42%) | 70 (40%) | 47 (47%) |  | 17 (6.5%) | 8 (4.4%) | 9 (12%) |  |
| **Wealth Category** |  |  |  | 0.034 |  |  |  | 0.058 |
| Low | 126 (45%) | 70 (40%) | 56 (55%) |  | 91 (35%) | 56 (31%) | 35 (45%) |  |
| Middle | 60 (22%) | 44 (25%) | 16 (16%) |  | 45 (17%) | 31 (17%) | 14 (18%) |  |
| High | 91 (33%) | 62 (35%) | 29 (29%) |  | 124 (48%) | 95 (52%) | 29 (37%) |  |
| **Binary Distance Group** |  |  |  | 0.048 |  |  |  | >0.9 |
| Far | 80 (29%) | 58 (33%) | 22 (22%) |  | 46 (18%) | 32 (18%) | 14 (18%) |  |
| Near | 197 (71%) | 118 (67%) | 79 (78%) |  | 214 (82%) | 150 (82%) | 64 (82%) |  |
| **Distance from Health Post** | 37.70 (31.81) | 41.44 (34.58) | 31.19 (25.14) | 0.016 | 30.75 (30.79) | 28.70 (28.53) | 35.51 (35.24) | 0.075 |
| **Wealth Index Raw Score** | -0.38 (0.24) | -0.36 (0.22) | -0.43 (0.25) | 0.002 | -0.28 (0.31) | -0.26 (0.31) | -0.33 (0.31) | 0.039 |

<sup>1</sup>n (%); Mean (SD)
<sup>2</sup>Pearson's Chi-squared test; Wilcoxon rank sum test

